# Reduction of brooding and more general depressive symptoms after fMRI neurofeedback targeting a melancholic functional-connectivity biomarker

**DOI:** 10.1101/2021.01.21.20248810

**Authors:** Jessica Elizabeth Taylor, Takashi Yamada, Takahiko Kawashima, Yuko Kobayashi, Yujiro Yoshihara, Jun Miyata, Toshiya Murai, Mitsuo Kawato, Tomokazu Motegi

## Abstract

Depressive disorders contribute heavily to global disease burden; This is possibly because patients are usually treated homogeneously, despite having heterogeneous symptoms with differing underlying neural mechanisms. On the contrary, treatment that directly influences the neural circuit relevant to an individual patient’s subset of symptoms might more precisely and thus effectively aid in the alleviation of their specific symptoms. We tested this hypothesis, using fMRI functional connectivity neurofeedback to target a neural biomarker that objectively relates to a specific subset (melancholic) of depressive symptoms and that is generalizable across independent cohorts of patients. The targeted biomarker was the functional connectivity between the left dorsolateral prefrontal cortex and left precuneus, which has been shown in a data-driven manner to be less anticorrelated in patients with melancholic depression than in healthy controls. We found that the more a participant normalized this biomarker, the more related (brooding and more general depressive), but not unrelated (trait anxiety), symptoms were reduced. Thus, one-to-one correspondence between a normalized neural network and decreased depressive symptoms was demonstrated. These results were found in two experiments that took place several years apart by different experimenters, indicating their reproducibility. Indicative of their potential clinical utility, effects remained one-two months later.

## Introduction

Depressive disorders contribute heavily to global disease burden (James et al. 2017, WHO 2020), however current treatments are of mediocre efficacy, with patients showing high rates of relapse and increased mortality risk (Paykel 2008, Lépine and Briley 2011). While depressive disorders are usually diagnosed by a medical doctor using interviews and questionnaires, a more objective alternative for diagnosis may be the use of a neuroimaging data-driven “biomarker” (Insel et al. 2010, Insel and Cuthbert 2015). Different symptoms of depression have differing underlying neural mechanisms (Lee and Kim 2014; Figure 1) and so patients with depression can be classified into different biotypes based on their resting-state fMRI functional connectivity (rs-FC; Williams 2016, Drysdale et al. 2017, Yahata et al. 2017, Tokuda et al. 2018, Yamashita et al. 2019, Kashiwagi et al. 2020, Yamashita et al 2020). Responsiveness to treatment may differ depending on ‘biotype’ (Williams 2016, Drysdale et al. 2017, Tokuda et al. 2018) rendering biomarker and biotype identification important and therapy which targets these pertinent for more advanced and precise treatment. Here, we describe a real-time fMRI neurofeedback paradigm which can be used to train the brains of people with depressive symptoms to function more like those of healthy people. Importantly, unlike traditional treatments, this method specifically targets a neural biomarker which has objectively been shown to relate to a specific subset (melancholic) of depressive symptoms and to be generalizable across independent cohorts of patients (Ichikawa et al. 2020).

**Figure 1.**
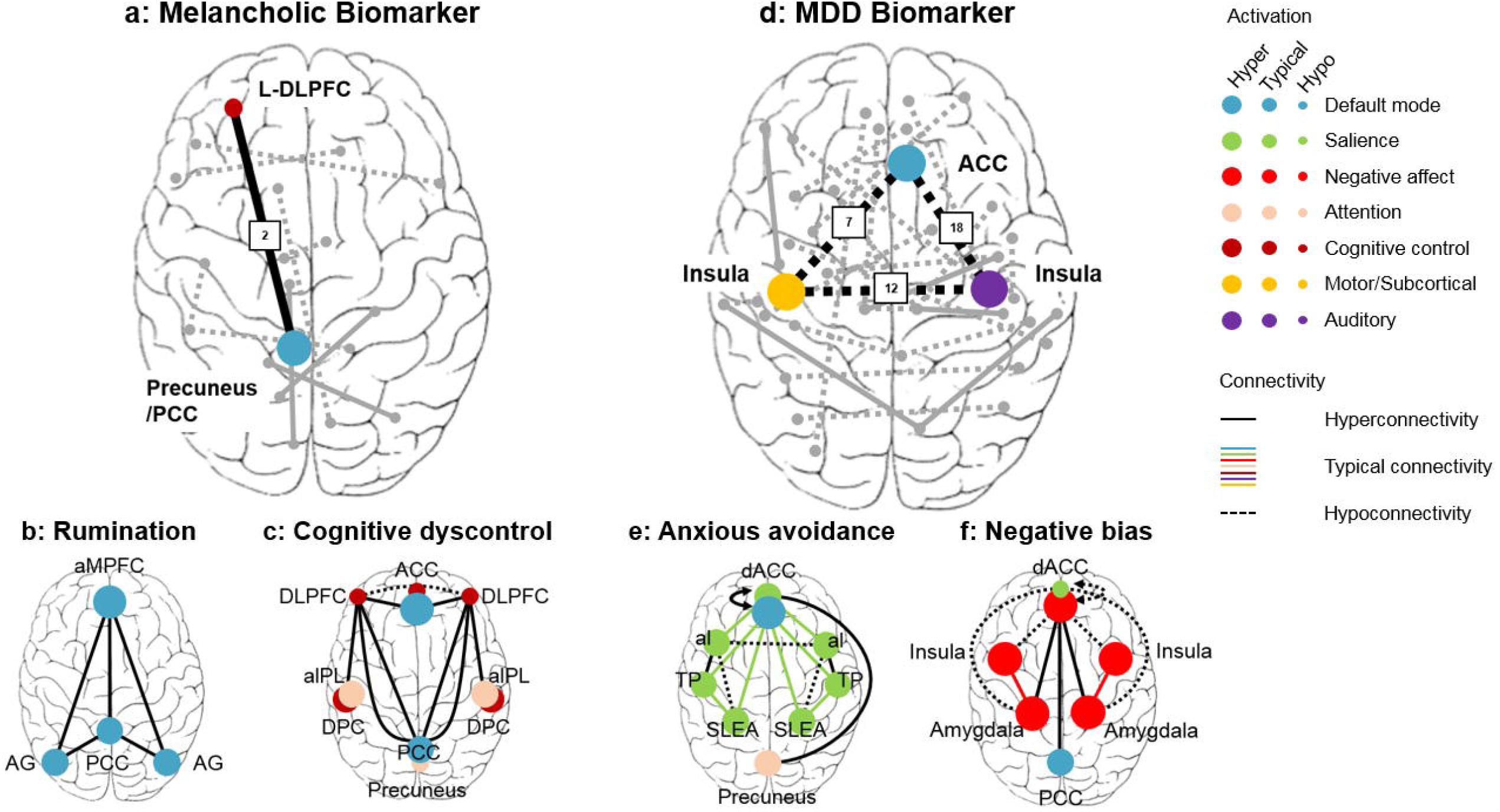
Altered FC between the DLPFC/mFG and precuneus/PCC is specific to the melancholic subtype of depression. Based on “biomarker” and “biotype” findings, which indicate that different neural networks underlie different subtypes of depressive symptoms, we hypothesized that FCNef targeting a biomarker for melancholic depression should lead to the alleviation of *mainly* melancholic symptoms. **a**. The Melancholic biomarker of Ichikawa et al. (2020). Hyperconnectivity between the left DLPFC/mFG and left precuneus/PCC contributed heavily to this biomarker (this FC is highlighted by the black line; other identified FCs are shown as grey lines); Hyperconnectivity between these regions was also shown in the ‘cognitive dyscontrol’ biotype network of Williams (2016), shown in c. **b**. The ‘rumination’ biotype proposed by Williams (2016), which involves FC disruptions within the default mode network that are anchored in the PCC. **c**. The ‘cognitive dyscontrol’ biotype proposed by Williams (2016). Just like the melancholic biomarker in a., this is characterized by hyperconnectivity between the left DLPFC of the cognitive control circuit and the PCC of the default mode. Note that this ‘cognitive dyscontrol’ biotype involves a loss of the usual anticorrelation between the DLPFC and the PCC; of which the PCC is part of the default mode network that is also disrupted in the ‘rumination’ biotype. Together a., b., and c. indicate that, during melancholic depression, prolonged activation of the PCC and resulting prolonged inactivation of the DLPFC causes FC between the DLPFC and PCC to become less anticorrelated than is found in healthy controls. In this situation of relative positive rather than anticorrelation between cognitive control and default mode networks, both cognitive and rumination symptoms may manifest. **d**. The MDD biomarker of Yamashita et al. (2020). Note that the identified FCs do not overlap with the FCs identified in the Melancholic biomarker shown in a. Of particular note, FC between the left DLPFC/mFG and left precuneus/PCC is not identified here, indicating that hyperconnectivity between these regions is specific to *melancholic* depression. These FCs identified here do, however, include hypoconnectivity between the ACC and bilateral insula (highlighted by black dotted lines), which is also a key region in the negative affect circuit evoked by negative emotion stimuli to characterize the ‘negative bias’ biotype of Williams (2016), shown here in f. Furthermore, hypoconnectivity between the left and right insula of the salience circuit, identified here (also highlighted by a black dotted line), is also a clinical phenotype that characterizes the ‘anxious avoidance’ biotype of Williams (2016), shown here in e. Other identified FCs are shown in grey. **e**. The ‘anxious avoidance’ biotype involving disruption of the salience circuit proposed by Williams (2016). Note that this does not overlap at all with the FCs identified in the Melancholic biomarker shown in a. Similar to the MDD biomarker shown in d., hypoconnectivity between the bilateral insula is identified here. **f**. The ‘negative bias’ biotype network, which involves disruption of the insula, ACC (and amygdala) within the negative affect circuit, proposed by Williams (2016). Note that this does not overlap at all with the FCs identified in the Melancholic biomarker shown in a. However, the hypoconnectivity between the ACC and bilateral insula proposed here matches well with that found in the MDD biomarker. Together d., e., and f. indicate that during the type of depression picked up by the MDD biomarker, hypoconnectivity between the bilateral insula may contribute to anxious symptoms and hypoconnectivity between the ACC and bilateral insula might contribute to negative bias symptoms. It is worth noting that MDD is not the same diagnosis as anxiety, although they are often comorbid. However, that is not problematic for this explanation since this is based on data-driven biotypes rather than clinical diagnoses. Overall, when a., b., and c., are compared with d., e., and f., it becomes clear that melancholic depressive symptoms have their own unique biotype, which includes hyperconnectivity between the DLPFC and precuneus/PCC. L-DLPFC=left dorsolateral prefrontal cortex. PCC=posterior parietal cortex (includes precuneus). aMPFC=anterior medial prefrontal cortex. AG=angular gyrus. ACC=anterior cingulate cortex. aIPL=anterior inferior parietal lobule. DPC=dorsal parietal cortex. dACC=dorsal anterior cingulate cortex. aI=anterior insula. TP=temporal pole. SLEA=sublenticular extended amygdala.

In neurofeedback participants are trained to modulate their own neural activity in order to influence their behavior and patterns of thinking. When the targeted regions of the brain are disease-related, such as a biomarker, then real-time fMRI neurofeedback may aid in the alleviation of psychiatric symptoms (Coben et al. 2010, Scheinost et al. 2013, Sulzer et al. 2013, Stoeckel et al. 2014, Sitaram et al. 2017, Yamada et al. 2017, Watanabe et al. 2017, Rance et al. 2018, Young et al. 2018, Lubianiker et al. 2019, Paret et al. 2019, Shibata et al. 2019, Tursic et al. 2020). While medicinal treatment of depression leads to relatively high relapse rates (Rush et al. 2006, Paykel 2008, Lépine and Briley 2011) and an array of unpleasant side-effects (Thase et al. 2005, Cascade et al. 2009), to date, no serious side-effects have been reported as arising as a consequence of real-time fMRI neurofeedback. This is potentially because, unlike medication, neurofeedback can be designed to specifically target only the disease-related regions of the brain. Furthermore, unlike psychological treatment (DeRubeis et al. 2005), neurofeedback efficacy should not depend much on the practitioner. If proven to be effective, therefore, real-time fMRI neurofeedback for depression may be of great aid in complementing or even replacing traditional treatments for depression. Several ‘proof of concept’ studies have already shown the potential effectiveness of real-time fMRI neurofeedback paradigms for depression (Linden et al. 2012, Young et al. 2014, 2017). Recently, a new type of real-time fMRI neurofeedback called functional connectivity neurofeedback (FCNef) has been developed (Koush et al. 2013, Megumi et al. 2015, Koush et al. 2017, Ramot et al. 2017, Yamada et al. 2017, Yamashita et al. 2017, Tsuchiyagaito et al. 2020), where participants are trained to modulate functional connectivity (FC) between selected regions of interest (ROIs). This type of neurofeedback has proven effective in changing the resting-state connectivity between intrinsic brain networks in the long-term (Megumi et al. 2015) and in leading to an improvement in aberrant behaviors related to neurobiological disorders (Ramot et al. 2017, Yamada et al. 2017, Tsuchiyagaito et al. 2020). FCNef, therefore, has high potential for the treatment of depressive symptoms.

For FCNef, the objective selection of an appropriate FC to target is of high importance. As indicated above, different FCs are likely to be more or less relevant depending on the subset of depressive symptoms to be treated. A specific subset of FCs relevant to *melancholic* depression were identified in a biomarker by Ichikawa et al. (2020; Figure 1a). Of these, FC between the left dorsolateral prefrontal cortex (DLPFC)/middle frontal gyrus (mFG) and the left precuneus/posterior parietal cortex (PCC) was shown to be of great importance; The DLPFC/mFG and precuneus/PCC belong respectively to the Executive Control Network (Seeley et al. 2007) and the Default Mode Network (Raichle et al. 2001), which reciprocally inhibit one another (Chen et al. 2013, Ferrier et al. 2020). This leads to alternations between a DLPFC/mFG-active precuneus/PCC-inhibited state and a precuneus/PCC-active DLPFC/mFG-inhibited state. Such fluctuations cause the FC between these regions to be anticorrelated in healthy people. However, if people spend an excess of time in the precuneus/PCC-active DLPFC/mFG-inhibited state (e.g. under situations of rumination and cognitive dyscontrol (Williams 2016), which are more common in melancholic depression (Nelson and Mazure 1985, Roca et al. 2015, Zaninotto et al. 2016; Figures 1b and 1c)), then fewer fluctuations occur. This results in a reduced anticorrelation (hyperconnectivity) between these regions, which was found in the aforementioned melancholic biomarker (Ichikawa et al. 2020). Training patients to increase the anticorrelation between these regions, via FCNef, might therefore alleviate melancholic depressive symptoms, including those of rumination. Interestingly, this FC was not identified as relevant in a generalizable biomarker for a more general diagnosis of depression (major depressive disorder (MDD); Yamashita et al. 2020; Figure 1d). This biomarker for MDD instead involves, among other perturbations, altered FC between the bilateral insula and between the bilateral insula and ACC; It thereby respectively overlaps with biotypes implicating clinical features of negative bias (Williams 2016; Figure 1f) and anxious avoidance (Williams 2016, Figure 1e). These results further support the idea that different neural networks dysfunctions give rise to different subsets of symptoms. FCNef that normalizes FC between the DLPFC/mFG and precuneus/PCC (makes it more like that of healthy controls) should therefore mainly only improve melancholic symptoms.

It should be noted that FC between the DLPFC/mFG-precuneus/PCC had the second greatest contribution to the biomarker of Ichikawa et al. (2020), however this was barely different from the FC with the greatest contribution. Interestingly, this FC has been identified as uniquely failing to become normalized via treatment with SSRIs (Ichikawa et al. 2020). Repetitive transcranial magnetic stimulation to one of the ROIs, the left DLPFC, has proven sometimes effective in the treatment of medication-resistant patients with depression (Richieri et al. 2017). Normalization of this particular FC with FCNef might therefore provide treatment for melancholic depression beyond that which can be achieved using medication.

We hypothesized that normalization of the DLPFC/mFG-precuneus/PCC FC should specifically predict decreases in melancholic symptoms. To investigate this and the reproducibility of effects, in two similar experiments we ran subclinical participants with depressive symptoms in a FCNef paradigm targeting this FC, rewarding them when it became more anticorrelated. Scores on the Beck Depression Inventory-II (BDI; Beck et al. 1996), and the Rumination Response Scale (RRS; Treynor et al. 2003, Hasegawa 2013) were measured before and after FCNef training. As a control, we also measured scores on the Trait Anxiety Scale (STAI2; Spielberger 1983), because anxiety has a different set of underlying neural dysfunctions (Lemche et al. 2016, Williams 2016; Figure 1e) and therefore should not be affected by changes in the FC we targeted. To further gauge the potential clinical effectiveness of this paradigm, we collected follow-up data for participants from the second experiment one- and two-months after they had completed the FCNef paradigm.

## Methods

### Ethics Statement

This study was approved by the Ethics Committee of the Review Board of Advanced Telecommunications Research Institute International, Japan, and by the Kyoto University Certified Review Board (UMIN000015249, jRCTs052180169). All experiments were performed in accordance with relevant guidelines and regulations. All participants provided written informed consent prior to participation.

### Participants

All participants were recruited based on screening questionnaires and clinician assessment. To meet criteria for participation in these experiments, participants must have (a) an average BDI score of over eight averaged across two BDI measurements (the range was 8.5-23.5, with a mean of 14.3 and a std of 5.1), (b) no inclination of suicidal thoughts, as measured by a question on the BDI, (c) no current or recent mental or psychiatric diseases, (d) understanding of the Japanese language. All participants had normal or corrected-to-normal vision. They were paid ¥8,000 for each MRI session (+ a bonus in some sessions, described below) and ¥3,000 for each questionnaire session (where they filled out the BDI, RRS and STAI2).

#### 1st FCNef Experiment

In total, nine participants (5 male, 4 female; 23.33±1.76 years old) participated in this experiment, which took place in 2016 and 2017. Specifically, these participants all completed the whole “fundamental experimental procedure”, which took place six days and was composed of the ‘functional localizer task’, SHAM, and FCNef Days 1-4 (explained in detail below). The BDI and the targeted rs-FC data (but not the RRS and STAI2 data) for seven of these participants have been reported elsewhere in a preliminary form (Yamada et al. 2017). The data for the other two participants was collected just after this previous publication by the same experimenter and so has been included in this data set.

#### 2nd FCNef Experiment

The design is basically the same as the 1st experiment, except for the additional examination of long-term effects. In total, 11 participants participated in this 2nd FCNef Experiment, which took place in 2019 and 2020, and was run by different experimenters from the 1st experiment. Specifically, these participants all completed the whole fundamental experimental procedure. The data from one participant was excluded, because (despite declaring no mental health problems when recruited) an in-depth interview with a psychiatrist revealed that she had just recovered from a strong case of MDD. This meant that the data of 10 participants (4 male, 6 female; 23.00 ±1.67 years old) was included in analyses. None had participated in the 1st FCNef experiment. Of these participants, nine came back for follow-up testing one-month after the main paradigm, and eight for follow-up testing two-months after the main paradigm.

### Materials

Visual stimulus presentation was controlled using MATLAB 7.5.0.342 (2007b; The MathWorks Inc.). The visual stimuli were projected to an opaque screen set inside the scanner via a (DLA-X7-B, JVC; frame rate = 60 Hz) projector and a mirror system. Participants responded to the stimuli using MRI compatible response pads (HHSC-2 × 2, Current Designs, Inc., PA, USA).

### Experimental procedure

For a general schematic of the experimental procedure, see Figure 2.

**Figure 2.**
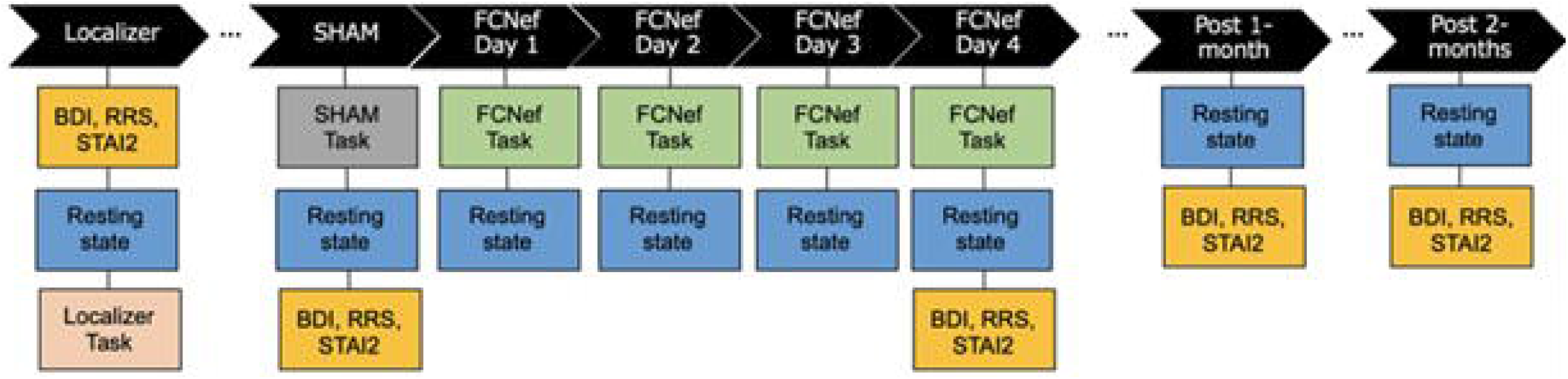
Experimental Outline. A schematic showing the general flow of experiments. Each vertical column represents a different experimental day. Periods are inserted between non-consecutive days. The post 1- and post 2-month data was only collected for the 2nd experiment. BDI = Beck’s Depression Inventory. RRS = Rumination Response Scale. STAI2 = Trait Anxiety Scale. SHAM = SHAM neurofeedback. FCNef = Functional Connectivity Neurofeedback.

#### Functional Localizer Task

The purpose of this task was to (a) identify peaks of activity within the left DLPFC/mFG and left precuneus/PCC under respective conditions where the Executive Control and Default Mode Networks are expected to be activated, and (b) use the identified peaks for each participant to make their individualized ROIs for FC calculation. Participants entered the scanner and their resting-state fMRI was taken. Here and in all other resting-state sessions this took 10 minutes and participants were simply instructed to relax and maintain a central fixation. After this, their T1-weighted structural MRI was taken and then the localizer task began. This was the famous ‘n-back’ task, which under difficult conditions requires recruitment of the Executive Control Network (Thompson et al. 2016); This allows us to identify peak DLPFC/mFG activity for each participant. On the contrary, when activity from the difficult conditions of this task are subtracted from activity during rest periods, then Default Mode Network activation is expected (Raichle et al. 2001); This allows us to identify peak precuneus/PCC activity for each participant. The specific details of this task, its analyses, and the building of individualized ROIs can be found in the Supplementary Materials. Example ROIs can be seen in Figure 3e and f.

**Figure 3.**
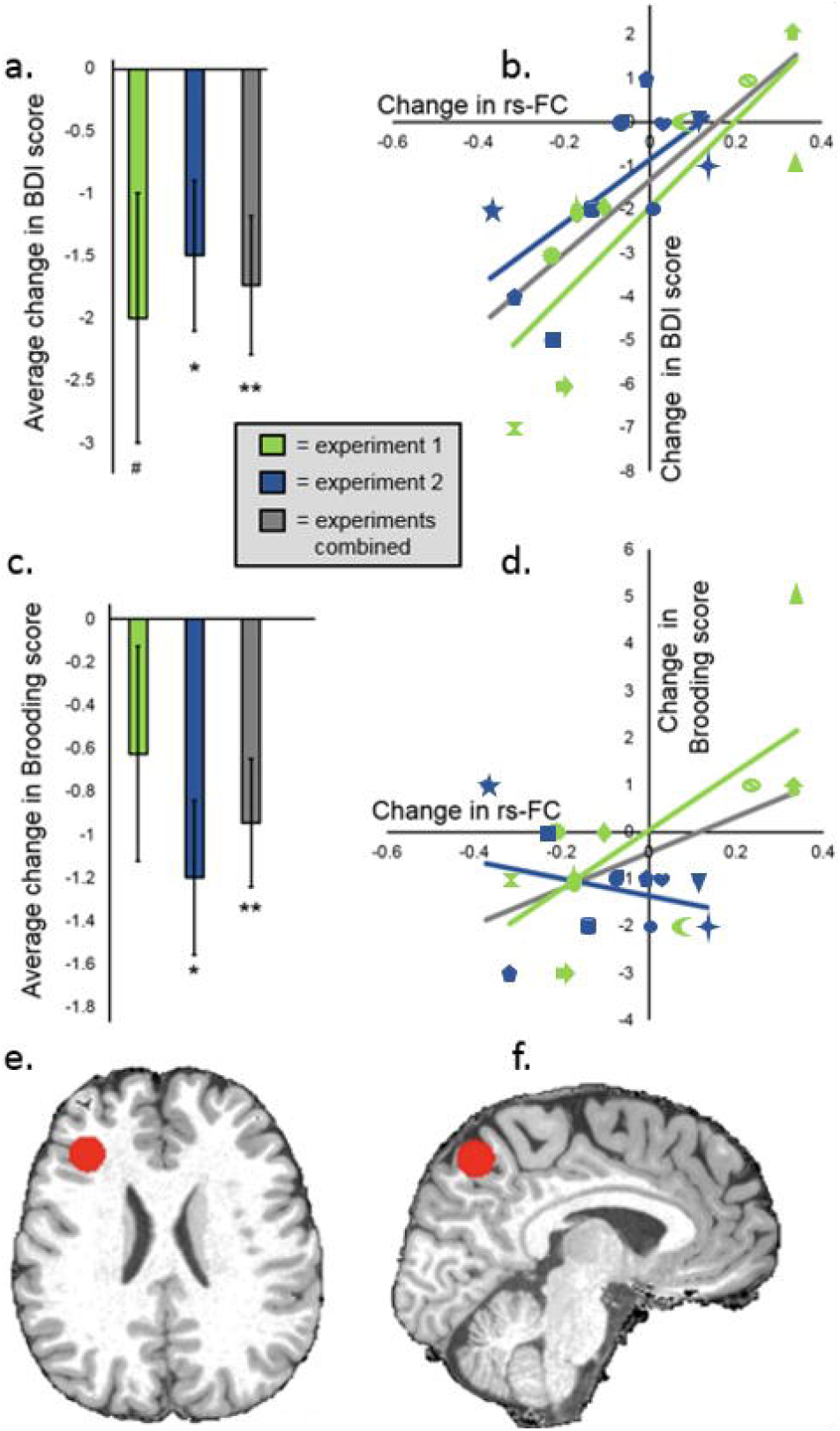
Changes that occurred in the initial period for BDI scores, RRS Brooding factor scores, and for rs-FC between the left DLPFC/mFG and left precuneus/PCC. All “changes” on this figure refer to those from SHAM to the last day of FCNef (data from SHAM was subtracted from that from FCNef Day 4). The individual shapes on b. and d. represent individual participants; The same shape is kept for each participant for data in b. and in d. **a**. Average reductions in BDI scores from SHAM to the last day of FCNef (for the 1st experiment t(8)=-2.00, p=0.08; for the 2nd experiment t(9)=-2.50, p=0.03; for the experiments combined t(18)=-3.12, p<0.01). **b**. Correlations between changes in BDI scores and changes in DLPFC/mFG-precuneus/PCC rs-FC. In both experiments, the more this rs-FC became normalized (i.e. the stronger the anticorrelation), the greater the reduction in a participant’s BDI scores (for the 1st experiment r=0.84, p<0.01; for the 2nd experiment r=0.67, p=0.03; for the experiments combined r=0.74, p<0.001). **c**. Average reductions in Brooding factor scores, from SHAM to the last day of FCNef, (for the 1st experiment t(8)=-1.26, p=0.25; for the 2nd experiment t(9)=-3.34, p<0.01; for the experiments combined t(17)=-3.18, p<0.01) **d**. Correlations between changes in Brooding factor scores and changes in DLPFC/mFG-precuneus/PCC rs-FC (for the 1st experiment r=0.69, p=0.04; for the 2nd experiment r=-0.28, p=0.42; for the experiments combined r=0.45, p=0.05) **e**. An example participant’s left DLPFC/MFG ROI that was made in their own subject-space is shown here, in red, rendered on their skull stripped T1. **f**. The same example participant’s left precuneus ROI that was made in their own subject-space is shown here, in red, rendered on their skull stripped T1. BDI = Beck’s Depression Inventory. Brooding = Rumination Response Scale’s Brooding factor. rs-FC = resting-state Functional Connectivity. SHAM = SHAM neurofeedback. FCNef = Functional Connectivity Neurofeedback.

#### SHAM

The purpose of this task was to calculate participants’ FC while they were doing the FCNef task without real feedback. This provides a baseline for each individual with which their FCs from FCNef can be compared. On this day participants entered the scanner and completed five sessions of SHAM FCNef, each with six trials. On each trial, during the ‘induction period’, participants had been instructed to try their best to “do something with their brain” to get the best feedback possible. During instructions, a list of example strategies had been provided, so that participants understood what was meant by “do something with your brain”. It is possible that this may have influenced what participants did. Nonetheless, participants were not told to use any explicit strategies on any given trial or session, meaning that they had to learn how to get favorable feedback via trial and error during the task. The strategies used in each session were reported to the experimenter. None of these (and none from the example list) related to depression or the n-back task. Feedback was calculated as a score (0-100) which was then presented as a circle on screen. Participants had been instructed that the larger this circle was in circumference, the better the feedback (minimum circumference reflects a score of 0, maximum circumference reflects a score of 100). They were informed that they would get a real cash bonus corresponding to the sum of feedback across trials, and that they should therefore do their best to get the best feedback by “making this circle as big as possible” on each trial. In reality, during SHAM FCNef, all feedback was random (so that the overall average score was 50). This allowed us to calculate each participants’ “baseline FC”. All participants received a cash bonus of ¥500. Further details about this task, its analysis, and how baseline FCs were calculated can be found in the Supplementary Materials. After participants had completed SHAM, their resting-state fMRI was taken. Finally, they filled out the BDI, RRS, and STAI2.

#### FCNef Days 1-4

On Day 1 participants received an apology and an explanation that feedback on the previous day (SHAM) had been random, but that it would be real from the current day onwards. On all days, participants entered the scanner and the FCNef task began. This task was completely identical to the SHAM task, except that feedback was really based on the participants’ neural activity. Again, participants were not told to use any explicit strategies on any given trial or session and no reported strategies were related to depression or the n-back task. On each trial, the FC between the participant’s individualized ROIs in the left DLPFC/mFG and left precuneus/PCC was calculated online and compared to their baseline FC (see above and the Supplementary Materials for details). The more negative the FC during the induction period was than that individual participant’s baseline, the higher the score and thus the bigger the feedback circle presented on that trial (up to mean − 1 standard deviation where the circle reached its maximum size). By using this feedback, which corresponds to a real cash bonus (¥500∼¥3,000), and by not explicitly providing participants with strategies to use, the goal was to implicitly reinforce (Ramot et al. 2017) a more negative FC (more in line with that of healthy people) between these ROIs for each of the participants. On Day 4, after exiting the scanner, participants filled out the BDI, RRS, and STAI2.

#### Follow-up (Post) testing (one- and two-months after FCNef)

Resting-state fMRI was taken. After this finished, participants filled out the BDI, RRS, and STAI2.

### Imaging data acquisition

A 3T scanner with a 32-channel head coil, located at the ATR Brain Activity Imaging Center, was used for scanning acquisition (Siemens MAGNETOM Verio, Siemens, Erlangen, Germany). Anatomical images were acquired using a T1-weighted MP-RAGE protocol (slice number, 240; matrix size, 256 * 256; FOV, 256 mm; voxel size, 1.0 * 1.0 * 1.0 mm (no slice gap); TR, 2300 ms; TE, 2.98 ms; flip angle, 9°). T2*-weighted images reflecting blood oxygen level-dependent (BOLD) signals were acquired in all experimental and resting state sessions using gradient-echo echo-planar imaging (EPI) (slice number, 60; matrix size, 100 * 100; FOV, 200 mm; voxel size, 2.0 * 2.0 * 2.0 mm (no slice gap); TR, 1000 ms; TE, 28 ms; flip angle, 65°). Each ‘functional localizer task’ session took 590s and consisted of 590 volumes. Each SHAM and FCNef session took 512s and consisted of 512 volumes. Each resting state session took 600s and consisted of 600 volumes. The first ten volumes taken in each session of all experimental and resting state sessions were discarded to ensure steady-state magnetization.

### Data analyses

Online calculation of feedback is described in the Supplementary Materials.

#### BDI, RRS, and STAI2 results

BDI and STAI2 scores were separately totaled for each participant individually, on every day they were measured. RRS scores were totaled for each participant for each of the three factors (Depression, Brooding, and Reflection) separately. “Changes” in scores were calculated by subtracting the score from the SHAM day from the score on a later day (FCNef Day 4, 1-month later, or 2-months later). The data from one outlier was excluded from the analysis of Brooding factor score change (the Brooding factor change for this participant = 5, whereas the overall mean (when this participant was included) was −0.63 ±1.82). The data from one outlier was excluded from the correlation calculated between BDI and targeted rs-FC changes from SHAM to 1-month later (the BDI change for this participant at this time = −6 and their rs-FC change was 0.25, whereas the overall means (when this participant was included) were −0.68±2.5 for BDI change and −0.09±0.21 for rs-FC change).

#### Calculating DLPFC/mFG-precuneus/PCC resting-state FC (rs-FC) offline

SPM8 (Wellcome Trust Centre for Neuroimaging, University College London, UK) was used to pre-process and analyze the imaging data. Standard pre-processing steps were completed in the following order: slice-timing correction, realignment, normalization to the skull stripped T1, and spatial smoothing using a Gaussian filter (FWHM = 6 mm). This data was then denoised via linear regression, with 6 motion parameters, a parameter for average signal over the whole brain, a parameter for average signal from cerebrospinal fluid, a parameter for average signal from grey matter, and parameters for the derivatives of all aforementioned parameters. It was scrubbed so that volumes with framewise displacement >0.5mm were removed (Power et al. 2014). A temporal bandpass filter was applied to the time series using a Butterworth filter with a pass band between 0.008 Hz and 0.1 Hz. The resulting time-series from the two ROIs were extracted and Pearson’s coefficient was then calculated between them. “Changes” in rs-FC between the targeted ROIs were calculated by subtracting the Pearson’s coefficient representing this rs-FC from the SHAM day from that on a later day (FCNef Day 4, 1-month later, or 2-months later). These “changes” in the targeted rs-FC were correlated with “changes” in BDI scores, with “changes” in scores on the three factors of the RRS, and with “changes” in STAI2 scores.

## Results

The FCNef task scores (reflecting neurofeedback success) significantly increased from the first to the last day of FCNef (Supplementary Table 3; t(18)=-2.31, p=0.03), indicating the success of the neurofeedback training. Unless specified otherwise, “change(s)” refers to results from SHAM subtracted from those on the final day of FCNef; They thereby reflect changes in results from before to after FCNef.

### Detailed Results

Participants’ average scores on the questionnaires and their rs-FC between the DLPFC/mFG-precuneus/PCC are shown for experiments individually and combined in Supplementary Table 1. The specific details for correlations taken between changes in the targeted rs-FC and changes in scores on the questionnaires are shown in Supplementary Table Daily FCNef task scores, and the relation between these and participants’ changes in scores on the questionnaires, are shown in Supplementary Table 3. Finally, in the Supplementary Materials, the results of the two experiments separately and combined are provided and results of statistical tests are given.

#### BDI and DLPFC/mFG-precuneus/PCC rs-FC

We ran a linear mixed-effects model (LME) with data from the two experiments combined (LME-1). This had a dependent variable of BDI change and an independent variable of targeted rs-FC (that between the DLPFC/mFG and precuneus/PCC) change. A likelihood ratio test showed that including a regressor for Experiment (1st or 2nd) and a regressor for its interaction with the targeted rs-FC change did not improve the model (‘BDI change∼ rs-FC change*Experiment’; AIC with the Experiment regressor and its interaction = 79.69; without them = 77.29; χ2(2)=3.60, p=0.17). This indicates that the results were not different for the two experiments. An ANOVA using the model excluding the regressors for Experiment and its interaction (‘BDI change∼ rs-FC change’) showed a main effect of targeted rs-FC change (f(1,17)=23.36, p<0.001). These results indicate that change in the targeted rs-FC, from SHAM to the last day of FCNef, was a significant predictor of change in BDI score. Regardless of whether the data of both experiments were analyzed separately or combined, a positive correlation between changes in the targeted rs-FC and BDI changes was found (Figure 3b). These results indicate that, regardless of experiment, as the targeted rs-FC became normalized, depressive symptoms (BDI scores) were reduced. Participants’ overall reduction in depressive symptoms proved significant (Figure 3a).

In order to exclude the possibility that the above results were obtained due to outlier participants, we excluded one participant from the model and re-ran it with the same regressors. Using the estimated coefficients for the targeted rs-FC change when this participant was excluded we estimated their BDI change. We repeated this so that each participant was left-out once, meaning that we had estimated BDI changes for each participant. We then correlated these with their real BDI changes (Figure 4). Estimated and real BDI changes correlated significantly and positively (r=0.65, p<0.005). These results indicate that LME-1 was not affected by outliers and therefore provide robust support for the idea that changes in rs-FC between the targeted ROIs from before to after FCNef are predictive of changes in depressive symptoms.

**Figure 4.**
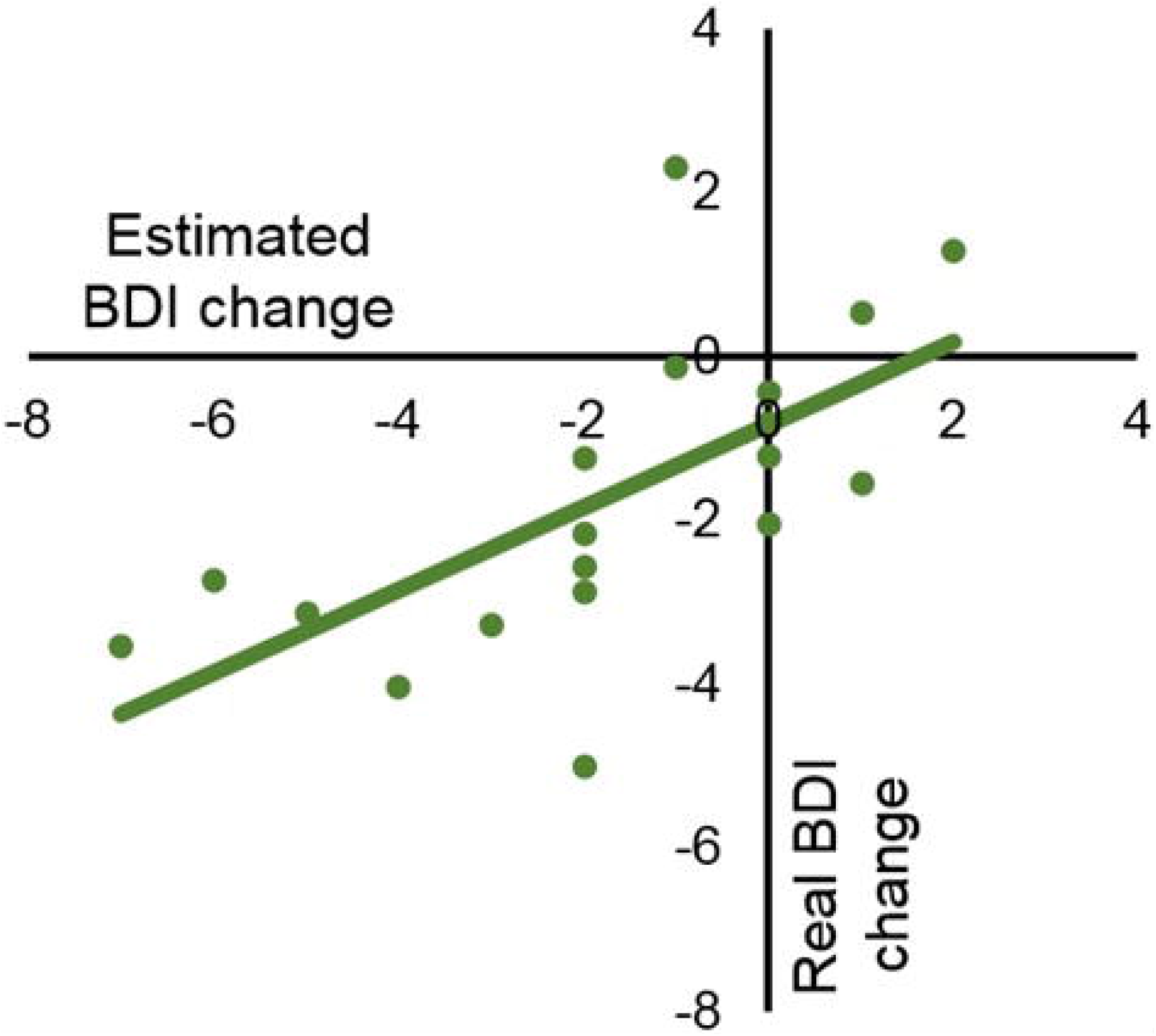
Correlations between real and estimated changes in BDI scores from before to after FCNef. LME-1 well explained participants’ changes in BDI scores from before to after FCNef using their changes in DLPFC/mFG-precuneus/PCC rs-FC over the same time-period. This supports the idea that, with FCNef, changes in this rs-FC lead to changes in depressive symptoms. To ensure the results of this model weren’t simply driven by outliers, we next calculated the coefficients for changes in this rs-FC with each participant left out. For each participant, their changes in BDI scores were then estimated using the coefficients from the model from which they were left out. The estimated BDI changes correlated significantly and positively with real BDI changes (r=0.65, p<0.005), showing that this model does not overfit and can well explain participants’ data. BDI = Beck’s Depression Inventory. rs-FC = resting-state Functional Connectivity. SHAM = SHAM neurofeedback. FCNef = Functional Connectivity Neurofeedback.

In the 2nd experiment, BDI scores and rs-FC between the targeted ROIs were followed up one- and two-months after participants had completed the main paradigm (detailed information is in the Supplementary Materials). Overall, BDI scores were found to remain lower than at SHAM at both time points, but not significantly so (Figure 5a). Correlations between changes in BDI score and the targeted rs-FC remained significant one-month later and were maintained in a similar direction even two-months later (Figure 5b). Longitudinal data was not collected for the 1st experiment and so a between-experiment comparison cannot be made for this.

**Figure 5.**
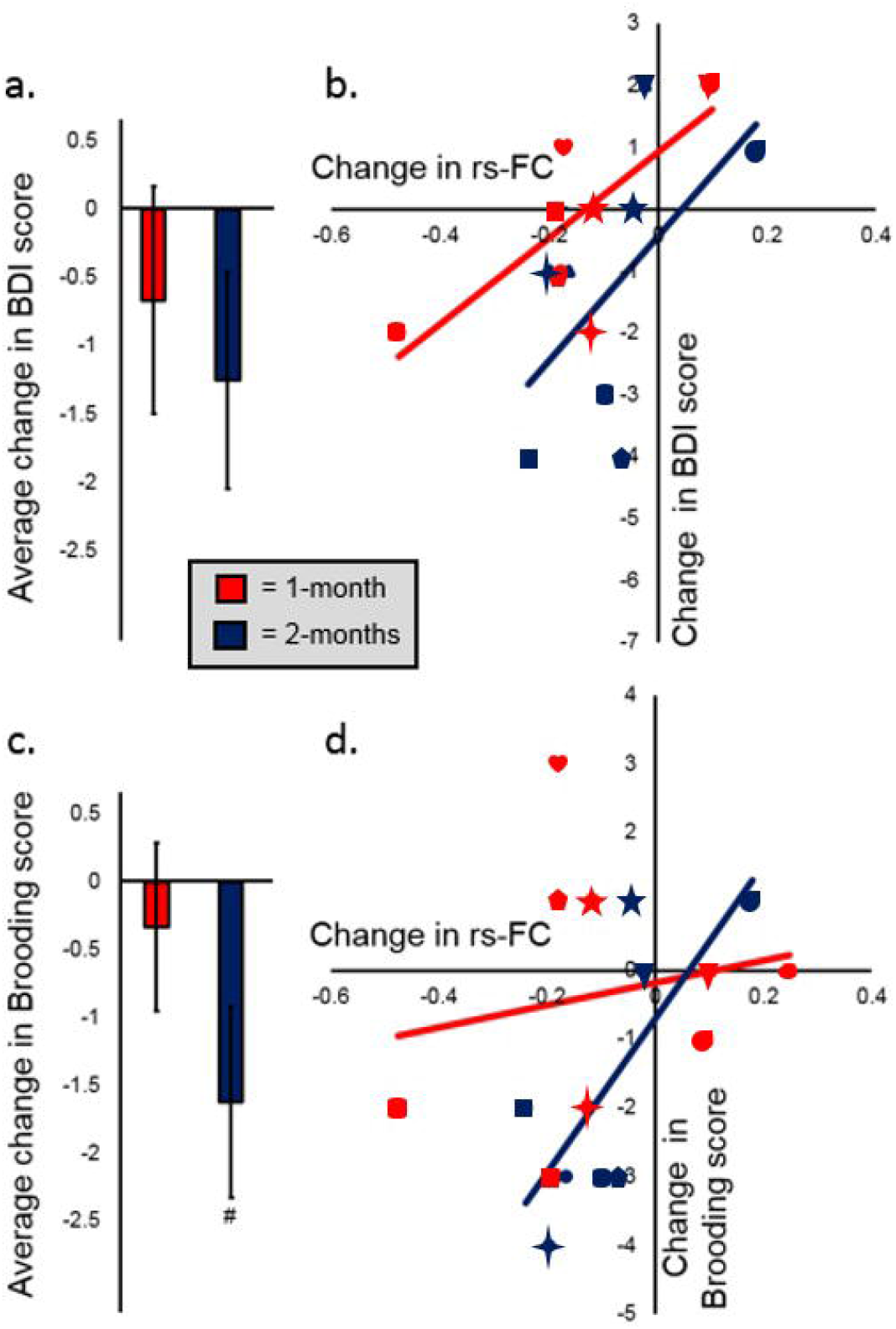
Changes that occurred in the longer term for BDI scores, RRS Brooding factor scores, and for rs-FC between the left DLPFC/mFG and left precuneus/PCC. Data was examined in the longer term only for the 2nd experiment. “Changes” on this figure refer either to those from SHAM to 1-month later (data from SHAM was subtracted from that from 1-month later; shown in red) or to those from SHAM to 2-months later (data from SHAM was subtracted from 2-months later; shown in dark blue). The individual shapes on b. and d. represent individual participants from the 2nd experiment; The shape for each individual participant shown in Figure 3 (2nd experiment) are maintained here and kept consistent for the the1- and 2-months data and for data in b. and in d. **a**. Average reductions in BDI scores from SHAM to 1- and 2-months after FCNef (for 1-month later t(8)=0.80, p=0.45; for 2-months later t(7)=1.570, p=0.16). **b**. Correlations between changes in BDI scores and changes in the targeted rs-FC. The more this rs-FC became normalized (i.e. the stronger the anticorrelation), the greater the reduction in a participant’s BDI scores; This was significant 1-month after SHAM (r=0.78, p=0.02) and still in the same direction 2-months after SHAM (r=0.58, p=0.13). **c**.Average reductions in Brooding factor scores from SHAM to 1- and 2-months after FCNef (for 1-month later t(8)=0.54, p=0.61; for 2-months later t(7)=2.30, p=0.06). **d**. Correlations between changes in Brooding factor scores and changes in the targeted rs-FC. The more this rs-FC became normalized (i.e. the stronger the anticorrelation), the greater the reduction in a participant’s Brooding factor scores. This was not significant 1-month after SHAM (r=0.18, p=0.64), but it was significant 2-months after SHAM (r=0.73, p=0.04). BDI = Beck’s Depression Inventory. Brooding= Rumination Response Scale’s Brooding factor. rs-FC = resting-state Functional Connectivity. SHAM = SHAM neurofeedback. FCNef = Functional Connectivity Neurofeedback.

#### RRS, STAI2, and DLPFC/mFG-precuneus/PCC rs-FC

The FC we targeted in FCNef was selected because it was part of a biomarker for *melancholic* depression; We therefore wished to examine how changes in this rs-FC related specifically to changes in melancholic symptoms of depression. The RRS can be separated into three factors: Depression, Brooding, and Reflection (Treynor et al. 2003). Of these factors, the Brooding factor is specifically thought to reflect maladaptive rumination (Treynor et al. 2003), which is thought to be a specific trait of melancholic depression (Nelson and Mazure 1985). A LME similar to LME-1 was run, but with changes in scores on the Brooding factor of the RRS (instead of BDI changes) as the dependent variable (LME-2; Brooding change∼ rs-FC change). A likelihood ratio test showed that including a regressor for Experiment (1st or 2nd) and a regressor for its interaction with changes in the targeted rs-FC did not significantly improve the model (Brooding change∼ rs-FC change*Experiment’; AIC with the Experiment regressor and its interaction = 63.24; without them = 63.58; χ2(2)=4.34, p=0.11). This indicates that the results were not different for the two experiments. An ANOVA using this model showed no significant main effects or interactions (ps>0.05). However, correlational findings showed a trend (p=0.05) towards significance indicating that, overall, as the targeted rs-FC became normalized, participants’ Brooding factor symptoms tended to be reduced (Figure 3d). Reductions in Brooding symptoms proved significant (Figure 3c).

In the 2nd experiment, participants’ RRS factor scores and their DLPFC/mFG-precuneus/PCC rs-FCs were followed up one- and two-months after participants had completed the main paradigm. Brooding scores changes at these time points are shown on Figure 5c. The relationship between changes in Brooding factor scores and changes in the targeted rs-FC went from completely unrelated when data from one-month later was compared to SHAM to being significantly related when data from two-months later was compared to SHAM (Figure 5d). Again, longitudinal data was not collected for the 1st experiment and so a between-experiment comparison cannot be made for this.

LMEs similar to those reported above were run, but with changes in scores on the RRS Depression factor, the RRS Reflection factor, and the STAI2 as dependent variables. Nothing of significance was found for any of these LMEs. This was regardless of whether factors of Experiment and its interaction were included or excluded. When long-term effects were investigated, correlations between changes in these scores and changes in the targeted rs-FC were not significant one- or two-months later (See the Supplementary Materials). Together, all aforementioned results indicate that the FC targeted is specifically related to melancholic depressive symptoms and not just overall sense of wellbeing (or lack thereof).

## Discussion

Depressive and brooding symptoms decreased after FCNef training targeting a biomarker for melancholic depression (FC between the left DLPFC/mFG and left precuneus/PCC). While these symptoms that are related to this biomarker correlated with its normalization, symptoms that are unrelated did not. These effects lasted into the long-term. Displaying their reproducibility and robustness; these results were found in two experiments which were carried out by different experimenters several years apart. Our FCNef paradigm may therefore provide reproducible results and thus be useful in targeted treatment of melancholic depression. These results also more generally indicate the potential validity of the technique of using FCNef to target data-driven, generalizable, biomarkers for psychiatric disease and/or subsets of symptoms (see also: Yahata et al. 2016, Takagi et al. 2017, Yoshihara et a. 2020).

Overall, general depressive (BDI) scores significantly decreased from before to after FCNef training (Figure 3a). Importantly, it was found that the degree to which the targeted FC became normalized was a significant and robust predictor of how much a participant’s BDI scores would reduce (Figure 3b, Figure 4). These results indicate that our FCNef paradigm has potential clinical relevance for the treatment of depressive symptoms. The reproducibility of these findings between experiments show their robustness and clearly supports the generalizability of the biomarker developed by Ichikawa et al. (2020). It was additionally found that the relationship between decreases in depressive scores and normalization of the targeted rs-FC was maintained significantly one-month later and in the same direction even two-months later (Figure 5b). To our best knowledge, this is the first study where the long-term effects of FCNef have been shown with clinical implications for depression (see also Ramot et al. 2017 for ASD). Although more expensive than medication, these long-term effects support the practical utility of FCNef for clinical interventions (Rance et al. 2018).

Overall, participants’ scores on the Brooding factor of the RRS significantly decreased after FCNef training (Figure 3c). These scores are thought to reflect maladaptive rumination (Treynor et al. 2003), which is common in melancholic depression (Roca et al. 2015). Brooding scores may have reduced here because we specifically targeted an FC which has a ROI that is highly related to rumination (Williams 2016, Misaki et al. 2020). Targeting the particular FC(s) related to the particular subset(s) of symptoms that a patient displays (and not those related to subsets of symptoms that they do not display) should allow for better neurofeedback precision. A recent study by Tsuchiyagaito et al. (2020) also found rumination scores to be reduced after FCNef. The targeted FC in this study was that between the temporoparietal junction and precuneus, both within DMN; it therefore shared one ROI (precuneus) with our own. However, in contrast to our own FC selection, which was made based on Ichikawa et al.’s biomarker made from whole-brain data-driven analyses, Tsuchiyagaito et al. selected their FC based on a connectome-wide search restricted to the medial PFC and PCC/precuneus.

In our study, decreases in brooding symptoms tended to correlate positively with normalization of the targeted FC (Figure 3d). In the 2nd experiment, this was additionally and significantly found for data over a longer-period (from SHAM to two months later) (Figure 5d) (see also long-term effects in Rance et al. 2018). Together these results indicate that changes in this rs-FC relate to changes in brooding symptoms. In total, results implicate FCNef targeting this FC as having potential clinical relevance in the treatment of the symptoms specifically related to this FC.

Overall, the finding that normalization of the targeted rs-FC related to decreases in depressive (BDI) and brooding (RRS) symptoms *from before to after* FCNef training is consistent with the idea that FCNef *caused* participants’ DLPFC/mFG-precuneus/PCC rs-FCs to normalize and thus their symptoms to reduce. However, because our analyses were based on correlational relationships, and because there was no control group, it is instead possible that symptoms spontaneously improved, resulting in the found changes in this rs-FC (Thibault et al. 2017, Sorger et al. 2019, Ros et al. 2020). For example, this result may have arisen due to a placebo effect, although no participants were told the purpose of this experiment and none used explicit strategies related to depression. Even in this case, our results support generalizability of the biomarker proposed by Ichikawa et al. (2020). Nonetheless, for the following reasons, it seems likely that the symptom changes found arose due to FCNef-induced changes in the targeted rs-FC. First, the feedback score significantly increased from the first to the last day of FCNef (Supplementary Results and Supplementary Table 3), which cannot be explained by a placebo effect. Second, we found decreases *only* in symptoms specifically related to our biomarker (BDI and Brooding scores). Trait anxiety symptoms, which are thought to be driven by different neural mechanisms (Lemche et al. 2016, Williams 2016; Figure 1), were initially found to be higher in our subclinical participants than those typically reported for healthy controls (see Supplementary Results and Donzuso et al. 2014). These were neither found to decrease with FCNef training nor to relate to changes in the targeted rs-FC. If our results occurred because our participants simply started feeling better, then these scores should also have decreased. Furthermore, the idea that our FCNef paradigm is what caused the found decreases in rs-FC and symptoms fits with a previously proposed model and empirical data (Shibata et al. 2019). Of course, although current results look promising, future study with our paradigm is still required before anything can be concluded about its causal effects.

Yamada et al. (2017) ran three therapy-resistant patients with MDD in this FCNef paradigm. The depressive symptoms of these patients were found to dramatically reduce, which is promising but still very preliminary. In the future, now that our results have demonstrated the safety and potential efficacy of this paradigm in subclinical participants, it should be further tested in clinical trials with real patients. This is one of the main objectives of Japanese Brain/MINDS Beyond project https://brainminds-beyond.jp/ and researchers from ATR, Kyoto University, and Hiroshima University are currently working on this.

In summary, normalization of the targeted rs-FC was found to correlate with decreases in related (ruminative brooding and more general depressive), but not unrelated (trait anxiety) symptoms. These effects were found to be reproducible over two experiments and to remain for at least one-two months later, indicating that they are robust and that the FCNef paradigm may have real clinical utility. Although further testing with a control group and with clinical patients are required, overall our results have proven promising for the treatment of depressive symptoms with our FCNef paradigm.

## Data Availability

The datasets generated and analysed during the current study are
not publicly available due to restrictions of their containing information that could compromise the privacy of research participants.
But the datasets are available from the corresponding author on reasonable request.

## Acknowledgements

We thank Kaori Nakamura for her help in scheduling and conducting the experiments; Taiki Oka, Misa Murakami, and Rumi Yorizawa for their help in screening participants; and Toshinori Yoshioka for his continued support with regards to the experimental scripts.

Furthermore, we would like to thank the team from the ATR Brain Activity Imaging Center (BAIC), in particular Akikazu Nishikido and Ichiro Fujimoto, for their technical support. Last, but not least, we would like to thank Leanne Maree Williams, Yasumasa Okamoto, Ayumu Yamashita, Naho Ichikawa, Go Okada, Toshinori Chiba, Aurelio Cortese, Tomohisa Asai, Yuto Kashiwagi, Okito Yamashita, Hiroshi Imamizu, Ryuta Tamano, Takeshi Ogawa, Motoaki Kawanabe, Junichiro Yoshimoto, Noriaki Yahata for their valuable comments on an earlier version of the manuscript.

