## Supplementary Materials for "Reduction of brooding and more general depressive symptoms after fMRI neurofeedback targeting a melancholic functional-connectivity biomarker"

**Methods**

**Functional Localizer Task.** Each participant completed three sessions of this task. At the beginning of each session, participants first had a rest-period, where they saw a black fixation cross which was presented on screen for 30s. They had been instructed to simply focus on this and relax. In each session, subsequent to the rest-period, there were eight blocks of an n-back- task, with the task ‘rule’ changing from block-to-block (order randomized within and between sessions). At the beginning of each block, written instructions were first presented on screen to inform participants of the current ‘rule’ and this was followed by 10 trials in which this ‘rule’ should be applied. On each trial, a fixation (for 1s) and then a number, between 1-9, (for 2s) was presented centrally on screen. In the ‘0-back’ blocks (of which there were two per session), the rule was to press the response button on every trial (every time a number appeared on screen). In the ‘1-back’, ‘2-back’, and ‘3-back’ blocks (each of which there were two per session), the rule was to press the response button on the current trial if the number that appeared on screen was the same as the number that had been presented on screen one, two, or three trials beforehand, respectively. A “rest” period (identical to the one at the beginning of the session) was inserted halfway through (between block 4 and 5 of) each session. Participants’ task was to follow the current ‘rule’ to make as many correct responses as possible. They performed at almost ceiling level.

**SHAM.** Participants each completed five sessions of SHAM FCNef. In each session of SHAM FCNef, there was first a 150 second rest (of which the first 10 scans were discarded), during which participants were simply instructed to relax and focus on the onscreen fixation cross. This was followed by six trials of SHAM FCNef. On each trial, the participant first saw a fixation cross with an equals sign above it for 14s. They had been instructed to simply maintain fixation on this fixation cross and not think about anything too deeply. Next, the equals sign above the fixation cross changed to a plus sign for 40s (this 40s is referred to as the ‘induction period’). Participants had been instructed that, while the plus sign was on screen, they should try their best to ‘do something with their brain’ to get the best feedback possible. They were never told an explicit strategy to use to try to do this on any given trial or session, and they were never recommended to maintain or switch strategies between trials or sessions. The experimenter simply asked which strategy they had used. It should be noted, however, that during instructions at the beginning, a list of example strategies had been provided so that participants had a better idea of what was meant by “doing something with their brain”. Participants sometimes used these strategies, and so were likely influenced somewhat by this list. Importantly for this experiment, none of the examples on the list were explicitly related to depression or the n-back task. After the 40s, unbeknownst to the participants (because the plus sign remained on screen) there was a 2s pause to allow for calculation of feedback. The feedback was subsequently presented on screen for 6s. Feedback was presented as a green circle with a fixation point in the center. For computation of the feedback during SHAM, the experimental script called upon the matlab 'rand' function and specified to use a Mersenne Twister generator for random number generation. It then used the ‘normrnd’ function to get a random number from a normal distribution with mean parameter 50 and standard deviation parameter 30.3.

**Data Analyses**

**Using the Functional Localizer Task to make individual ROIs.** The classifier for melancholic depression created by Ichikawa et al. (2020) was made based on averaged data from 130 individuals and the rs-FCs were calculated based on ROIs identified using anatomical parcellation. The results of this paper are therefore very telling in terms of overall regions of the brain that function differently for individuals with and without melancholic depression. However, they might not be appropriate for targeting using FCNef. This is because they are larger than ROIs generally targeted in FCNef and it is unlikely that these whole anatomically parcelled ROIs will activate fully for all individuals. Instead, smaller subsections within these larger ROIs are likely to be recruited, with the exact location of these activities differing somewhat between subjects. Indeed, this is what we found when we inspected our participants’ neural activity from the functional localizer task. Because we specifically wished to target the parts of this biomarker that participants actually use, we therefore used anatomically parcelled left DLPFC/mFG and left precuneus ROIs as a guideline and identified- for each participant individually- smaller subregions within these that were active when the Executive Control and Default Mode networks, respectively, were expected to be recruited. ROIs were made for each participant based on these subregions. The FC between these individually identified smaller ROIs was then targeted with FCNef. Because we found overall results that look promising for our FCNef paradigm, this technique of using a data-driven biomarker to determine the general region and then using a functional localizer to determine participant-specific regions might be useful for determining target ROIs for other neurofeedback paradigms in the future.

SPM8 (Wellcome Trust Centre for Neuroimaging, University College London, UK) was used to pre-process and analyze the imaging data. Standard pre-processing steps were completed in the following order: slice-timing correction, realignment, normalization to the skull stripped T1, and spatial smoothing using a Gaussian filter (FWHM = 4 mm). Left mFG and left precuneus masks (from the Automated Anatomical Labelling (AAL) atlas (Tzourio-Mazoyer et al. 2002); were also normalized (via inverse deformation) to the skull stripped T1. A whole-brain first level factorial model was made for each participant with the activity from all three sessions of their ‘functional localizer task’ combined. Activity from the different types of blocks (0-back, 1-back, 2-back, or rest) was modelled as different conditions. For each subject, the general linear model was used to fit the [fMRI](https://www.sciencedirect.com/topics/neuroscience/functional-magnetic-resonance-imaging) time series. Each condition was modeled from the onset until the offset of the relevant blocks. The six motion parameters were included as regressors of no interest. Once each participant’s model had been estimated, the following t-contrasts were estimated: 2-back>rest and rest>1-back+2-back. The Executive Control network is expected to be more active during the task than during rest (Thompson et al. 2016) and so the 2-back>rest contrast should reveal each participant's activity during executive control. On the other hand, the Default Mode Network is expected to be more active during rest than during this task (Raichle et al. 2001) and so the rest>1-back+2-back contrast was used to reveal each participant’s activity when this network should have been active. For each participant, their 2-back>rest contrast was masked with their subject-space normalized left mFG mask and the peak of activation (p<0.05 FWE corrected, minimum voxel size =10) within this was determined. Likewise, for each participant their rest>1-back+2-back contrast was masked with their subject-space normalized left precuneus mask and the peak of activation (p<0.05 FWE corrected, minimum voxel size =10) within this was determined. Subsequently, MarsBar (Brett et al. 2002) and the individual participants’ T1s were used to build ROIs with radiuses of 8mm for each participant centered around these peaks. This resulted in two ROIs for each person in their own individualized brain space- one for the left mFG and one for the left precuneus. These ROIs were subsequently used as targets for each participant for calculating their feedback online during FCNef and for analysis of their related rs-FC.

**Calculating ‘baseline’ FC from SHAM.** Code from the FCNef toolbox (available from https://bicr.atr.jp/decnefpro/software/) and fMRI data from SHAM were first used offline to determine a ‘baseline’ FC for each participant. In brief, SPM8 (Wellcome Trust Centre for Neuroimaging, University College London, UK) was used to realign and reslice volumes from the SHAM fMRI time-series to a reference volume (which itself had been realigned to fit with the data from the ‘functional localizer task’). This data was then denoised via linear regression, with six motion parameters, a parameter for average signal over the whole brain, a parameter for average signal from cerebrospinal fluid, a parameter for average signal from grey matter, and parameters for the derivatives of all aforementioned parameters. If there were any volumes with framewise displacement >0.5mm then this was added as a regressor as well (Power 2014). The data was then filtered using a Butterworth filter (with a pass band between 0.008 Hz and 0.3 Hz). Next, correlation coefficients close to, but not exactly the same as, Pearson’s coefficients were calculated using the signal from the two ROIs. During this calculation, to better baseline the signal, the mean signal from each ROI from the rest period (instead of from the induction period, which, if used, would have made this was a pure Pearson’s coefficient calculation) was subtracted from the signal from the same ROI from the induction period. The resulting coefficients were then transformed into a Fisher’s z coefficients. The average and standard deviations of these coefficients was determined and then these were transformed back to correlation coefficients. The resulting average correlation coefficient for each participant was used as their ‘baseline’.

**Calculating online feedback during FCNef.** For each participant, volumes from each trial were realigned and resliced online to a reference volume (which itself had been realigned to fit with the data from the ‘functional localizer task’) using SPM8 (Wellcome Trust Centre for Neuroimaging, University College London, UK). At the end of each induction period, volumes from that trial were denoised via linear regression and filtered, with the same regressors and parameters as were used when determining the baseline. The resulting time-series from each ROI were then correlated using the same method as above, to get a correlation coefficient similar to a Pearson’s correlation coefficient. This correlation coefficient was compared to the participant’s ‘baseline’ FC and converted into a score (0 = ‘baseline’ FC + one standard deviation or more; 50 = ‘baseline’ FC; 100 = ‘baseline’ FC - one standard deviation or more). This score was then used to determine the circumference of the feedback circle which was shown on screen. All of this online processing was conducted using code from the FCNef toolbox (available from https://bicr.atr.jp/decnefpro/software/).

**Results**

**1st FCNef Experiment**

**Results from FCNef Day 4 compared to SHAM.** For the nine participants in this experiment, average BDI scores decreased from SHAM to the last day of FCNef (FCNef Day 4), although this only reached a trend for significance (t(8)=2.00 p=0.08). RRS scores for all three factors (Brooding, Depression, and Deflection) did not significantly change (ps>0.05) and neither did STAI2 scores (t(8)=0.48 p=0.65). The rs-FC of interest decreased from SHAM to the last day of FCNef, although not significantly (p>0.05). These questionnaire scores and participants’ rs-FCs can be seen on Supplementary Table 1,

In support of the idea that this FC is related to depressive symptoms, a significant correlation was found between decreases in BDI and decreases in the targeted rs-FC from SHAM to the last day of FCNef; the more negative this rs-FC became the more participants’ depressive symptoms were reduced (r=0.84, p<0.01). In support of the idea that this FC is related to specifically *melancholic* depressive symptoms, a significant correlation was found between decreases in the Brooding factor of the RRS and decreases in this rs-FC from SHAM to the last day of FCNef; the more negative this rs-FC became the more participants’ depressive symptoms were reduced (r=0.69, p=0.04). Decreases in the other two factors of the RRS (Depression and Reflection) and decreases in STAI2 did not relate to decreases in this rs-FC from SHAM to the last day of FCNef (ps>0.05).

**2nd FCNef Experiment:**

**Results from FCNef Day 4 compared to SHAM.** For the 10 participants in this experiment, BDI scores significantly decreased from SHAM to the last day of FCNef (t(9)=2.50, p=0.03). Scores from the Brooding factor (t(9)=3.34, p<0.01), but not the Depression or Reflection factors of the RRS (ps>0,05) were significantly decreased from SHAM to the last day of FCNef. STAI2 scores did not significantly change from SHAM (t(9)=-1.45 p=0.18). Similar to results found in the 1st FCNef experiment, the rs-FC of interest also decreased from SHAM to the last day of FCNef, although not significantly (p>0.05). These questionnaire scores and participants’ rs-FCs can be seen on Supplementary Table 1

Interestingly, the correlation found in the 1st FCNef experiment between the decrease in BDI and the decrease in this rs-FC from SHAM to the last day of FCNef was replicated (r=0.67, p=0.03). The correlation found in the 1st FCNef experiment between the decrease in the Brooding factor of the RRS and the decrease in this rs-FC from SHAM to the last day of FCNef was not replicated for this time period (although see long-term results, below). No significant correlations between changes in scores for the other RRS factors or STAI2 and changes in the targeted rs-FC were found.

**Long-term Results**. BDI scores, RRS scores, STAI2 scores, and rs-FC between the DLPFC/mFG and precuneus/PCC were subsequently re-examined one-month after FCNef training for nine of the 10 participants and two-months after for eight of the 10 participants. For the nine participants who came back one-month later, BDI scores trended towards a significant reduction from SHAM (9.89±2.12) to the last day of FCNef (8.40±1.83; t(8)=2.16, p=0.06), and remained lower than at SHAM, but not significantly so, one-month later (9.22±2.07; t(8)=0.80,p=0.45). Scores from the RRS Brooding factor were significantly reduced from SHAM (9.44±1.13) to the last day of FCNef (8.33±0.93; t(8)=2.86, p=0.02), and remained lower than at SHAM, but not significantly so, one-month later (9.11±1.18; t(8)=0.54, p=0.61). Scores on the other two RRS factors did not significantly change (ps>0.05; for Depression they went from 23.00±2.28 to 21.33±2.32 to 20.33±2.72; for Reflection they went from 9.25±1.56 to 9.33±1.76 to 8.22±1.66). Likewise, scores on the STAI2 did not significantly change from SHAM (46.56±2.97) to the last day of FCNef (45.00±3.72; t(9)=-1.19, p=0.27) or to one-month later (44.89±3.03; t(8)=-1.38, p=0.20). The targeted rs-FC reduced from SHAM (0.043±0.48) to the last day of FCNef (-0.055±0.071), but not significantly (t(8)=1.61, p=0.15), and remained lower, but not significantly so, one-month later (-0.048±0.056; t(8)=1.29, p=0.23). For the eight participants who came back 1- and two-months later, BDI scores trended towards a significant reduction from SHAM (10.63±2.89) to the last day of FCNef (9.00±2.35; t(7)=2.23, p=0.06), and remained lower than at SHAM, but not significantly so, one- (9.75±2.40; t(7)=0.96, p=0.37) and two-months (9.38±1.99; t(7)=1.57, p=0.16) later. Scores from the RRS Brooding factor significantly reduced from SHAM (9.00±1.20) to the last day of FCNef (7.88±0.99; t(7)=2.55, p=0.03). They remained lower than at SHAM, but not significantly, so one-month later (8.25±1.25; t(7)=1.43, p=0.20) and trended towards being significantly lower than at SHAM two-months later (7.38±0.63; t(7)=2.30, p=0.06). Scores on the other two RRS factors did not significantly change (ps>0.05; for Depression they went from 21.13±2.88 to 20.50±2.45 to 19.63±2.98 to 18.63±1.65; for Reflection they went from 9.50±1.77 to 9.38±2.00 to 8.38±1.87to 8.38±1.38). STAI2 scores did not significantly decrease from SHAM (46.25±3.35) to the last day of FCNef (45.13±4.21; t(7)=-0.81, p=0.45), nor to one-month later (44.63±3.42; t(7)=-1.19, p=0.27), and nor to two-months later (44.00±3.36; t(7)=-1.45, p=0.19). Participants’ DLPFC/mFG-precuneus/PCC rs-FCs reduced, but not significantly so, from SHAM (-0.009±0.051) to the last day of FCNef (-0.122±0.076; t(7)=1.70, p=0.13), and remained lower than at SHAM, but not significantly so, one-month (-0.089±0.060; t(7)=1.02, p=0.34) and two-months later (-0.090±0.035; t(7)=1.78, p=0.12).

Correlations between changes in BDI score and FCNef were significant one-month later (r=0.78, p=0.02) and were maintained in a similar direction even two-months later (r=0.58, p=0.13). The relationship between changes between Brooding factor scores from the RRS and changes in the targeted rs-FC, went from completely unrelated when data from day 4 (r=-0.28, p=0.42) and one-month later (r=0.18, p=0.64) was compared to SHAM, to being significantly related when data from two-months later was compared to SHAM (r=0.73, p=0.04). Changes in the other two RRS factors (Depression and Reflection) and in STAI2 scores did not significantly correlate with changes in the targeted rs-FC one- or two-months later (ps>0.05).

**Experiments combined:**

**Results from FCNef Day 4 compared to SHAM.** For all participants combined, BDI scores on the last day of FCNef were found to have significantly decreased from SHAM (t(18)=-3.12, p<0.01). Scores from the Brooding factor (t(17)=-3.18, p<0.01), but not the factors of the RRS Depression (t(18)=-0.84 p=0.41) or Reflection (t(18)=-1.02 p=0.32) were found to have significantly decreased from SHAM. STAI2 scores did not significantly change from SHAM until the last day of FCNef (t(18)=0.52 p=0.61). The rs-FC of interest did not significantly decrease from SHAM to the last day of FCNef (t(18)=-1.00 p=0.33). These questionnaire scores and participants’ rs-FCs can be seen on Supplementary Table 1

The correlation between the decrease in BDI and the decrease in the targeted rs-FC from SHAM to the last day of FCNef was positive and significant (r=0.74, p<0.001). The correlation between the decrease in scores on the Brooding factor of the RRS and the decrease in this rs-FC from SHAM to the last day of FCNef was positive and trended towards significance (r=0.45, p=0.05). No significant correlations between changes in scores for the other RRS factors or STAI2 and changes in the targeted rs-FC were found. See Supplementary Table 2.

|  | **BDI Score** | | **RRS Brooding Score** | | **RRS Depression Score** | | **RRS Reflection Score** | | **STAI2** | | **rs-FC** | |
| --- | --- | --- | --- | --- | --- | --- | --- | --- | --- | --- | --- | --- |
|  | PRE | POST | PRE | POST | PRE | POST | PRE | POST | PRE | POST | PRE | POST |
| 1st Expt | 12.67  ±2.69 | 10.67  ±2.07 | 12.75  ±1.38 | 12.13  ±1.39 | 27.56  ±2.43 | 27.00  ±2.69 | 8.33  ±1.12 | 8.00  ±1.03 | 52.44  ±3.34 | 53.22  ±3.36 | -0.09  ±0.06 | -0.10  ±0.08 |
| 2nd Expt | 9.90  ±2.12 | 8.40  ±1.83 | 9.40  ±1.02 | 8.20  ±0.89 | 22.70  ±2.42 | 21.20  ±2.08 | 10.00  ±1.47 | 9.70  ±1.62 | 46.70  ±2.66 | 45.00  ±3.32 | 0.02  ±0.06 | -0.07  ±0.08 |
| Expts  Combined | 11.21  ±1.71 | 9.47  ±1.39 | 10.89  ±0.90 | 9.94  ±0.90 | 25.00  ±1.76 | 23.94  ±1.77 | 9.21  ±0.93 | 8.89  ±0.98 | 49.42  ±2.16 | 48.89  ±2.49 | -0.04  ±0.05 | -0.08  ±0.06 |

**Supplementary Table 1. The average and standard error of participants’ rs-FC and scores on the BDI, RRS, and STAI2 prior to and after FCNef training.** Pre = SHAM day (questionnaires and rs-FC was measured before the SHAM task). POST = FCNef Day 4 (questionnaires and rs-FC was measured after FCNef training on this day). Expt = Experiment. BDI = Beck’s Depression Inventory. RRS = Rumination Response Scale. STAI2 = Trait Anxiety Scale. rs-FC = resting-state Functional Connectivity

|  | | **BDI** | **RRS Depression** | **RRS Brooding** | **RRS Reflection** | **STAI2** |
| --- | --- | --- | --- | --- | --- | --- |
| rs-FC | SHAM | r=0.46  p=0.85 | r=0.23  p=0.34 | r=0.15  p=0.54 | r=0.08  p=0.74 | r=0.12  p=0.61 |
|  | FCNef  Day 4 | r=-0.36  p=0.12 | r=0.06  p=0.80 | r=0.18  p=0.46 | r=0.14  p=0.56 | r=-0.17  p=0.48 |
|  | Differences | r=0.74  p<0.001 | r=-0.17  p=0.94 | r=0.45  p=0.05 | r=0.32  p=0.18 | r=0.04  p=0.87 |

**Supplementary Table 2. Correlations between resting-state functional connectivity (rs-FS) and scores on the questionnaires.** The data from both experiments is combined here. The first row shows correlations between the targeted rs-FC and questionnaire scores on the SHAM day; the second row shows correlations between the targeted rs-FC and questionnaire scores on the final day of FCNef day (D4=day 4); the third row shows correlations between differences in rs-FC (FCNef D4- SHAM) and differences in questionnaire scores (FCNef D4- SHAM). BDI = Beck’s Depression Inventory. RRS = Rumination Response Scale. STAI2 = Trait Anxiety Scale. rs-FC = resting-state Functional Connectivity. SHAM = SHAM neurofeedback. FCNef = Functional Connectivity Neurofeedback.

|  | **Average FCNef task scores** | **Corr. between scores and BDI dif.** | **Corr. between scores and RRS Depression factor dif.** | **Corr. between scores and RRS Brooding factor dif.** | **Corr. between scores and RRS Reflection factor dif.** | **Corr. between scores and STAI2 dif.** |
| --- | --- | --- | --- | --- | --- | --- |
| SHAM | 52.24 ± 1.11 | r=-0.19  p=0.44 | r=-0.43  p=0.07 | r=0.02  p=0.95 | r=0.25  p=0.30 | r=-0.19  p=0.45 |
| FCNef Day 1 | 51.18 ± 3.27 | r=-0.01  p=0.97 | r=-0.14  p=0.57 | r=0.06  p=0.81 | r=-0.28  p=0.24 | r=-0.07  p=0.30 |
| FCNef Day 2 | 50.03 ± 3.91 | r=0.24  p=0.32 | r=0.07  p=0.77 | r=0.29  p=0.23 | r=0.12  p=0.61 | r=-0.04  p=0.88 |
| FCNef Day 3 | 59.13 ± 3.56 | r=-0.09  p=0.73 | r=0.21  p=0.40 | r=0.04  p=0.87 | r=-0.19  p=0.43 | r=0.29  p=0.23 |
| FCNef Day 4 | 61.36 ± 4.91 | r=0.02  p=0.93 | r=0.31  p=0.19 | r=0.32  p=0.19 | r=0.08  p=0.75 | r=0.20  p=0.42 |

**Supplementary Table 3. Participants’ average scores on the FCNef task each day and the correlations between these scores and differences in scores on the questionnaires (those from final day of FCNef - those on SHAM).** The data from both experiments is combined here. The first column shows the average and standard deviation of FCNef task scores shown on each day of FCNef training. Each subsequent column shows the correlation statistics between FCNef task scores presented to participants on each day and their difference (“dif”) in scores on the questionnaires from before to after FCNef. Because the questionnaires were not taken on every day of FCNef, we were unable to correlate daily scores on the task with daily scores on the questionnaires. Scores = scores on the FCNef task. Dif. = questionnaire scores from FCNef Day4 - those from SHAM. BDI = Beck’s Depression Inventory. RRS = Rumination Response Scale. STAI2 = Trait Anxiety Scale. rs-FC = resting-state Functional Connectivity. SHAM = SHAM neurofeedback. FCNef = Functional Connectivity Neurofeedback.
